# Biallelic variants in *MRPL49* cause variable clinical presentations, including sensorineural hearing loss, leukodystrophy, and ovarian insufficiency

**DOI:** 10.1101/2024.10.10.24315152

**Authors:** Huw B. Thomas, Leigh A.M. Demain, Alfredo Cabrera-Orefice, Isabelle Schrauwen, Hanan E. Shamseldin, Alessandro Rea, Thashi Bharadwaj, Thomas B. Smith, Monika Oláhová, Kyle Thompson, Langping He, Namanpreet Kaur, Anju Shukla, Musaad Abukhalid, Muhammad Ansar, Sakina Rehman, Saima Riazuddin, Firdous Abdulwahab, Janine M. Smith, Zornitza Stark, Samantha Carrera, Wyatt W. Yue, Kevin J. Munro, Fowzan S. Alkuraya, Peter Jamieson, Zubair M. Ahmed, Suzanne M. Leal, Robert W. Taylor, Ilka Wittig, Raymond T. O’Keefe, William G. Newman

**Author notes:** Correspondence to WG Newman, Manchester Centre for Genomic Medicine, Manchester University NHS Foundation Trust, Manchester M13 9WL, UK, or RT O’Keefe, Michael Smith Building, University of Manchester, Oxford Rd Manchester M13 9PT UK. These authors contributed equally (Huw B. Thomas, Leigh A.M. Demain, Alfredo Cabrera-Orefice, Ilka Wittig, Raymond T. O’Keefe, William G. Newman).

## Abstract

Combined oxidative phosphorylation deficiency (COXPD) is a rare multisystem disorder which is clinically and genetically heterogeneous. Genome sequencing identified biallelic *MRPL49* variants in individuals from five unrelated families with presentations ranging from Perrault syndrome (primary ovarian insufficiency and sensorineural hearing loss) to severe childhood onset of leukodystrophy, learning disability, microcephaly and retinal dystrophy. Complexome profiling of fibroblasts from affected individuals revealed reduced levels of the small and, a more pronounced reduction of, the large mitochondrial ribosomal subunits. There was no evidence of altered mitoribosomal assembly. The reductions in levels of OXPHOS enzyme complexes I and IV are consistent with a form of COXPD associated with biallelic *MRPL49* variants, expanding the understanding of how disruption of the mitochondrial ribosomal large subunit results in multi-system phenotypes.

---

The mitochondrial ribosome (mitoribosome) is a 55S ribonucleoprotein complex composed of large and small subunits, which co-ordinates the synthesis of the 13 proteins coded by the mitochondrial genome. These 13 proteins are vital components of the oxidative phosphorylation (OXPHOS) enzyme complexes. Human mitoribosomes are tethered to the mitochondrial inner membrane through the 39S large subunit of the mitoribosome^1^. This large subunit (mt-LSU) is comprised of two structural RNA molecules, a 16S ribosomal RNA (rRNA) and mt-tRNA^Val^, and 52 proteins^2–5^. In contrast, the 28S small subunit is comprised of 30 proteins and a 12S rRNA^6^. Several human diseases are caused by germline variants in genes encoding mitoribosome proteins^5,7^. However, despite the number of mitoribosome large subunit proteins, biallelic pathogenic variants have been identified in genes encoding only a limited number (Supplemental Table S1). Affected individuals with pathogenic variants in mt-LSU genes have a broad range of clinical presentations encompassing cardiomyopathy, liver dysfunction, neurological phenotypes and ovarian insufficiency^8–12^. In all reported individuals with disorders due to variants in genes encoding the mt-LSU, with the exception of *MRPL50*, there were measurable defects in mitochondrial OXPHOS, consistent with the definition of combined oxidative phosphorylation deficiency (COXPD, MIM 609060) disorders.

Here, we report on individuals from five unrelated families with biallelic variants in *MRPL49* characterized by diverse clinical phenotypes encompassing bilateral sensorineural hearing loss (SNHL) and primary ovarian insufficiency (POI, Perrault syndrome MIM 233400), microcephaly, learning disability, developmental delay, leukodystrophy, and retinal disease. Informed consent for diagnostic and research studies was obtained for all subjects in accordance with the Declaration of Helsinki protocols and approved by local institutional review boards.

A young South Asian woman and her sibling (Family 1 (F1)) were referred with a possible diagnosis of Perrault syndrome. The proband was an adult with bilateral, high frequency profound SNHL (Supplemental Figure S1), initially diagnosed at <5 years of age. She had absent menarche. Biochemical analysis revealed raised gonadotrophins consistent with hypergonadotropic hypogonadism. Pelvic imaging revealed absent ovaries, vagina, uterus and cervix. She was of normal height and weight but noted to have microcephaly (occipital frontal circumference [OFC] 50.6cm, -3.5 SD) and non-progressive mild intellectual disability. Her sibling was diagnosed with SNHL aged <5 years. She also has mild learning and behavioral problems and microcephaly (OFC 48.7cm, -3.7 SD). She has normal height and weight parameters and raised gonadotrophins consistent with hypergonadotropic hypogonadism. Pelvic imaging revealed a small uterus and absent ovaries (Supplemental Figure S3). She has been commenced on estradiol.

Exome sequencing did not identify variants in any of the known Perrault syndrome genes^13,14^ or genes associated with SNHL or POI in these two affected individuals. Autozygosity mapping using SNP arrays identified 11 homozygous regions >2Mb.

Within the largest region of autozygosity on chromosome 11 (∼50Mb), a rare homozygous variant in *MRPL49* c.275A>C, p.(His92Pro) (NM_004927 Figure 2A) was identified in both siblings. The variant was heterozygous in both unaffected parents and in a clinically unaffected sibling. Through Genematcher^15^ and international collaboration, we identified four additional families, including individuals presenting with biallelic variants in *MRPL49* displaying variable clinical presentations.

In F2, the same missense variant as identified in F1, *MRPL49* c.275A>C, p.(His92Pro), was identified in two siblings from a consanguineous family affected by intellectual disability and facial dysmorphism. It was not possible to formally assess the hearing status as the affected individuals were unable to undergo testing but there was no evidence of a hearing deficit. The female had no overt reproductive abnormalities.

The affected male had night blindness, bone abnormalities, borderline short stature (−1.8 SD) and seizures. He had borderline microcephaly at the time of initial assessment, but his current head circumference is within normal range. This family was also of South Asian ancestry and haplotype analysis of exomes from affected individuals in F1 and F2 revealed a shared haplotype of 1.24 Mb (chr11-64343482-65583893, GRCh38), indicating a common founder variant.

In a consanguineous South Asian family (F3) a homozygous c.262C>T, p.(Arg88Cys) variant in *MRPL49* was identified in a 5-10 year old boy with hearing loss. He could speak only monosyllables. He had episodes of fever associated with loss of consciousness treated with anticonvulsants. At 5-10 years of age, his weight was 12.54 kg (−2.6 SD), height was 103 cm with contractures (−1.6 SD) and head circumference was 47 cm (−3.2 SD). He had a small forehead, plagiocephaly, low anterior hairline, metopic prominence, strabismus, and orofacial dystonia. He exhibited hypertonia (lower limbs > upper limbs) and tremors.

In F4, an woman was identified in whom trio genome sequencing revealed a compound heterozygous missense variant c.262C>T, p.(Arg88Cys) *in trans* to a predicted loss of function frameshift variant c.125_126delTG, p.(Val42Glyfs*2) in *MRPL49*. She was referred as an adult for clinical genetics assessment to consider an underlying genetic cause for her intellectual disability, epilepsy, retinitis pigmentosa, bilateral posterior subcapsular cataracts, obesity, hypothyroidism treated with thyroxine, menstrual cycle anomalies and recent psychosis with neurocognitive decline. There was no history of SNHL. She has no known family history of similar clinical features. The affected individual’s clinical condition continued to deteriorate resulting in her death.

Of note, in an adult female with intellectual disability, bilateral SNHL requiring cochlear implants, kyphoscoliosis, and POI, a previous study had suggested that the molecular explanation was a homozygous variant in *PANX1*^16^. She also was homozygous for the same p.(Arg88Cys) *MRPL49* missense variant as in F3 and F4, which is likely to explain her clinical presentation (personal communication, Dale Caird).

In a consanguineous Middle Eastern family (F5) a different missense variant in *MRPL49* at the same 88 residue, c.263G>A p.(Arg88His), was identified in two siblings affected with developmental delay, SNHL, and ataxia. The profound hearing loss in the younger affected sibling (F5:II-2) was apparent after birth with distortion product otoacoustic emissions absent at all tested frequencies bilaterally. Brainstem auditory evoked potentials were abnormal with prolonged peripheral latencies and poor waveform reproducibility and poor proximal wave III/V with an increased threshold. He has never walked. At age 5-10 years, he has significant cognitive delay and is only able to say two words. His OFC is 46 cm (−4.1 SD). He had raised plasma lactate on multiple occasions. Throughout childhood he has experienced recurrent episodes of hypoglycemia. Recent assessment of renal function demonstrated urea of 7.9 mmol/L (at the upper end of the normal range 2.1 to 8.5), serum creatinine 50 umol/L (range 18-46), and raised serum potassium of 5.6 mmol/L (normal 3.6-5.2).

The elder affected sibling (F5:II-1) was more severely affected with a similar pattern of profound hearing loss and significant developmental delay. Ophthalmological assessment was unremarkable. His lactate was raised at 2.30 mmol/l. At age 5-10 years, he had stage 3 chronic kidney disease of unknown etiology (eGFR 51 ml/min/1.73m^2^). Renal ultrasound revealed slightly echogenic kidneys with mild distension of the left renal collecting system. He has recurrent hyperkalemia and a renal biopsy was consistent with chronic interstitial nephritis.

Brain magnetic resonance (MR) imaging was available from six affected individuals from the five families (Figure 1 and Supplemental figure S3). This imaging demonstrates consistent progressive patterns with symmetrical involvement of the globi pallidi in all six affected individuals, presenting as high T2 signal with or without associated diffusion restriction or as cystic change. In four individuals there is symmetrical T2 high signal in the deep white matter (centrum semiovale) with periventricular sparing. Involvement of the brain stem is seen in three and cerebellar atrophy in four of the six affected individuals. Globus pallidus cystic change, cerebellar atrophy and deep white matter T2 high signal changes were associated with more advanced disease.

**Figure 1:**
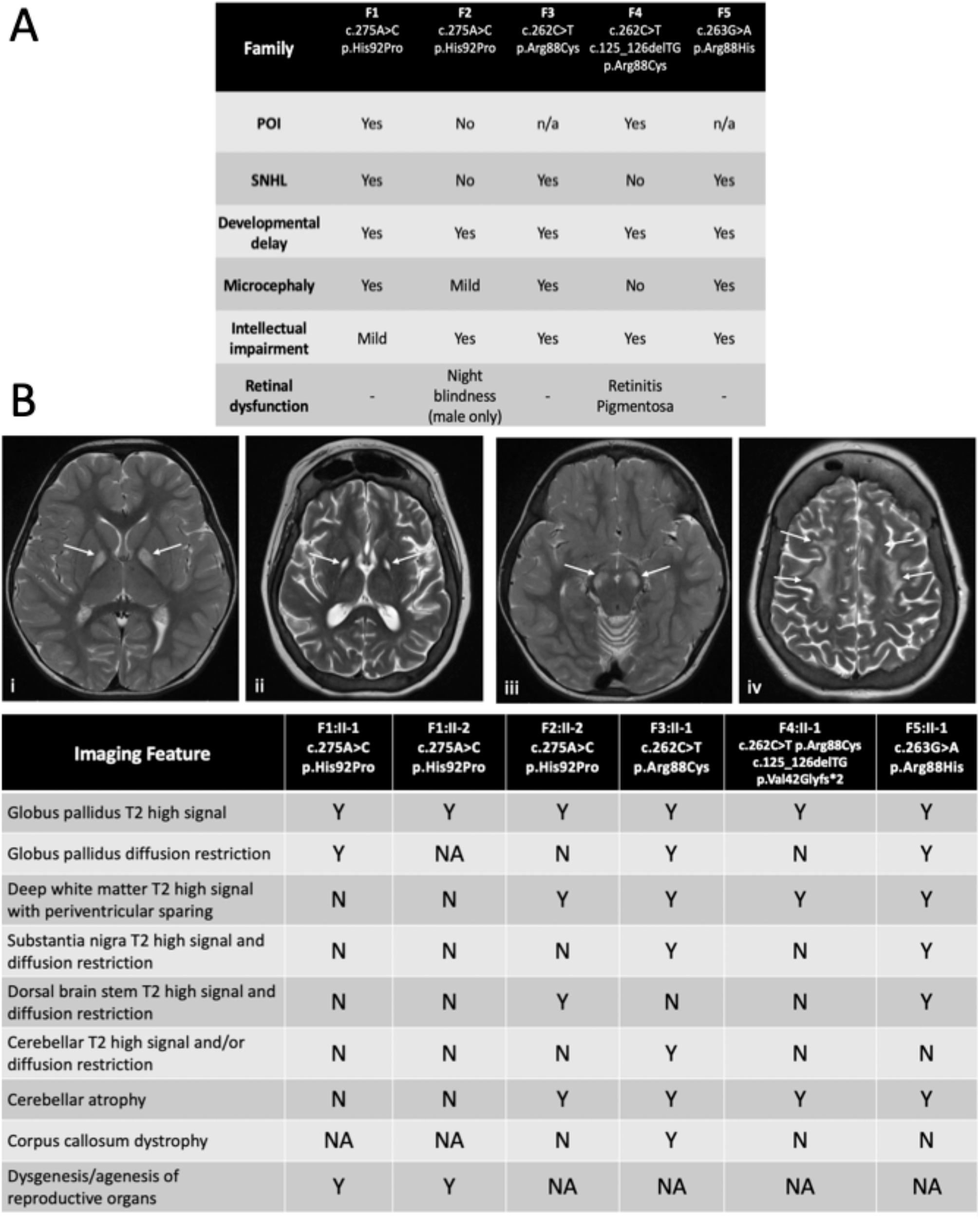
Clinical information and brain imaging data for families with biallelic *MRPL49* variants. **(A)**Clinical details of POI, SNHL and developmental/intellectual development for five unrelated families of individuals with either homozygous or compound heterozygous variants in *MRPL49*. **(B)**Selected axial brain MR images demonstrating features of patients with variants in *MRPL49*, and accompanying summary of all imaging features shared across 6 individuals from 5 unrelated families. All 6 affected individuals with available brain imaging had symmetrical T2 high signal changes in the globi pallidi (i). This feature progressed to symmetrical cystic changes in two affected individuals with more advanced disease (ii). In 3/6 affected individuals there was symmetrical T2 high signals and diffusion restriction (not pictured) involving the brain stem, including the substantia nigra (iii) and the dorsal brainstem. In 4/6 affected individuals there were symmetrical white matter T2 high signal changes, becoming more diffuse and confluent with more advanced disease (iv). There was also evidence of cerebral atrophy in 4/6 affected individuals, particularly involving the cerebellum (not pictured), but also affecting cortical grey matter and sub-cortical and deep white matter in individuals with more advanced disease (ii & iv).

To summarize the clinical features of the affected individuals with *MRPL49* biallelic variants (Figure 1), the majority of affected individuals have hearing loss (5/8), POI in post pubertal females (3/4), brain white matter changes (6/6), learning disability (8/8) and microcephaly (4/6). Additionally, some individuals experienced hypoglycemia, and had evidence of renal or retinal disease. The different biallelic variants in *MRPL49* in multiple unrelated families with similar clinical features meet the evidence criteria for a rare disease-gene association^17^. We proceeded to generate cellular and *in vitro* functional data to support this disease-gene association.

The amino acids in MRPL49 at positions 88 and 92, are conserved across multiple species (Figure 2B). In gnomAD v4.0 the variant alleles are reported at frequencies consistent with a rare monogenic disorder (Supplemental Table S2); of note His92Pro is represented as 22 of 86250 alleles in a South Asian subset^18^. No homozygous loss of function alleles are reported in gnomAD v4.0 and *in silico* predictors indicated that the missense variants would have a deleterious effect on function (Supplemental Table S3). Structurally residues Arg88 and His92 are situated at a beta-hairpin loop of the MRPL49 protein, which forms part of the protein interface with 16S rRNA (Figure 2C). Arg88 is a strictly invariant amino acid among all orthologs, forming an important network of hydrogen bonds with nearby residues. His92 is 95% conserved among orthologs, and packs against a hydrophobic core (Ile57, Pro60, Trp72). Substitutions of His92Pro, introducing a cyclic amino acid, as well as Arg88Cys and Arg88His (removing positive charge), are predicted to disturb the conformation of the MRPL49 beta-hairpin loop which could impact on interaction with the 16S rRNA and stability of the mt-LSU central protuberance.

**Figure 2:**
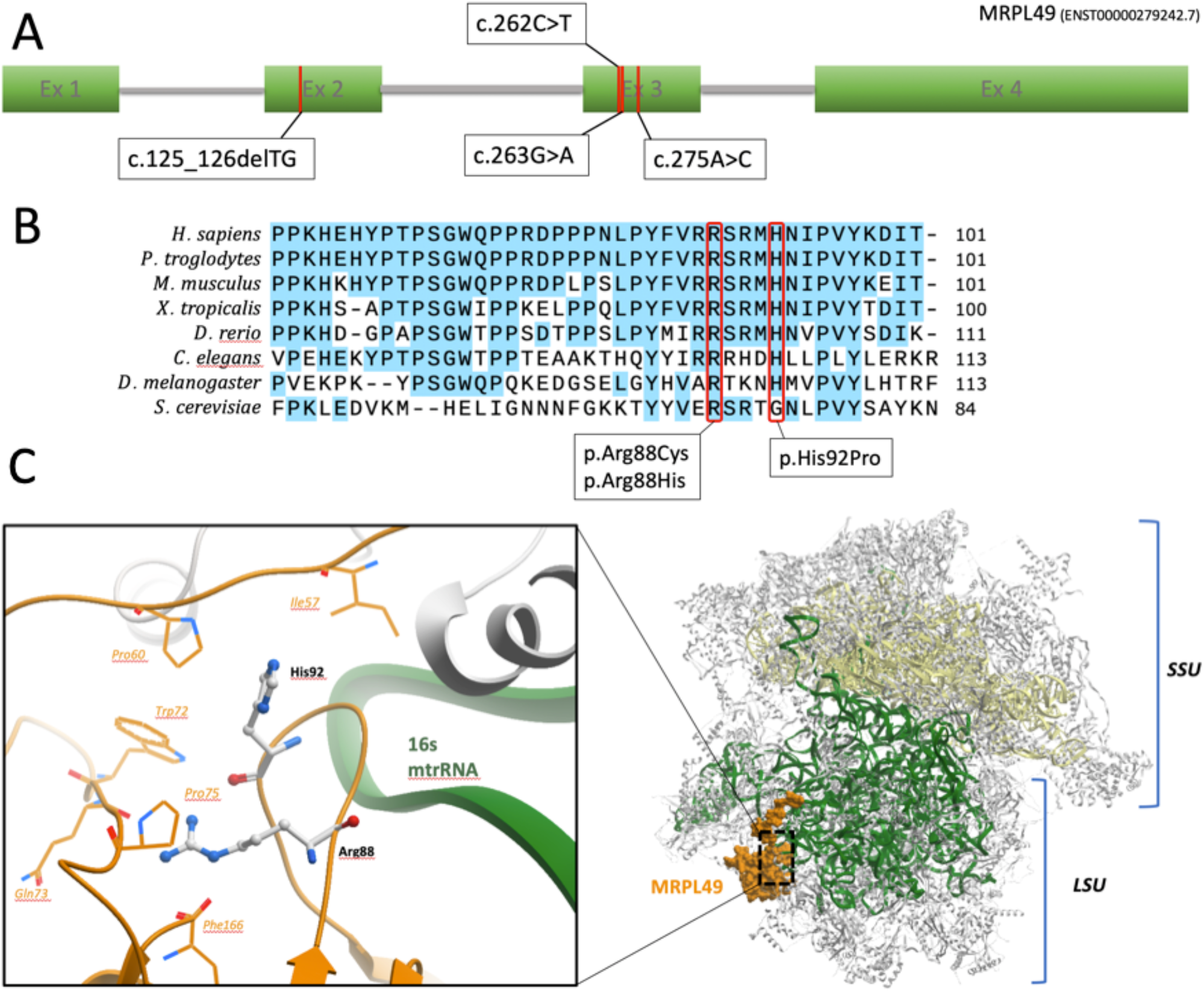
*In silico* modelling of MRPL49 variants. **(A)** Schematic representation of *MRPL49* transcript and disease-associated variant locations. All three missense variants are clustered in exon 3, whereas the single frameshift variant is located in exon 2. **(B)** Evolutionary conservation of MRPL49 affected residues across a broad range of orthologous species. Conserved residues are colored blue, variant amino acids (Arginine 88 and Histidine 92) are highlighted in red. **(C)** Three-dimensional representation of the location of MRPL49 (orange) within the human mitoribosome (PDB code:7QI4). Large (mt-LSU) and small (mt-SSU) colored in grey, 16S rRNA colored in green and 12S rRNA in yellow.

To consider this predicted disturbed interaction, we measured the 12S:16S rRNA ratio in dermal fibroblasts available from affected individuals in F1 and F5. Previous studies investigating disease-associated variants in genes encoding proteins of the mt-LSU have consistently demonstrated a relative reduction in the levels of 16S rRNA^10,19,20^. The levels of 16S rRNA were significantly reduced in the cells from both individuals with *MRPL49* variants compared to the 16S rRNA levels in control fibroblasts (p<0.0001 and p=0.0327, Figure 3A). These data demonstrate that the missense variants reduce 16S rRNA levels, consistent with the mechanism of other mt-LSU associated disorders and may impact upon translation of mitochondrially encoded proteins. To examine mitochondrial protein translation, we measured the activity and levels of components of the OXPHOS complexes in fibroblasts from affected individuals compared to control fibroblasts. OXPHOS enzyme analysis^21^ revealed decreased activity of a complex I subunit in both affected individuals (Figure 3B). Western blot analysis revealed reduced steady state levels of both complex I and complex IV subunits in fibroblasts from F5:II-1, but no significant changes in cells from F1:II-1. These differences reflect the sensitivity of the assays, as evidenced by the subsequent complexome profiling data, but the greater reductions in F5 are consistent with the more severe clinical phenotype in this affected individual.

**Figure 3:**
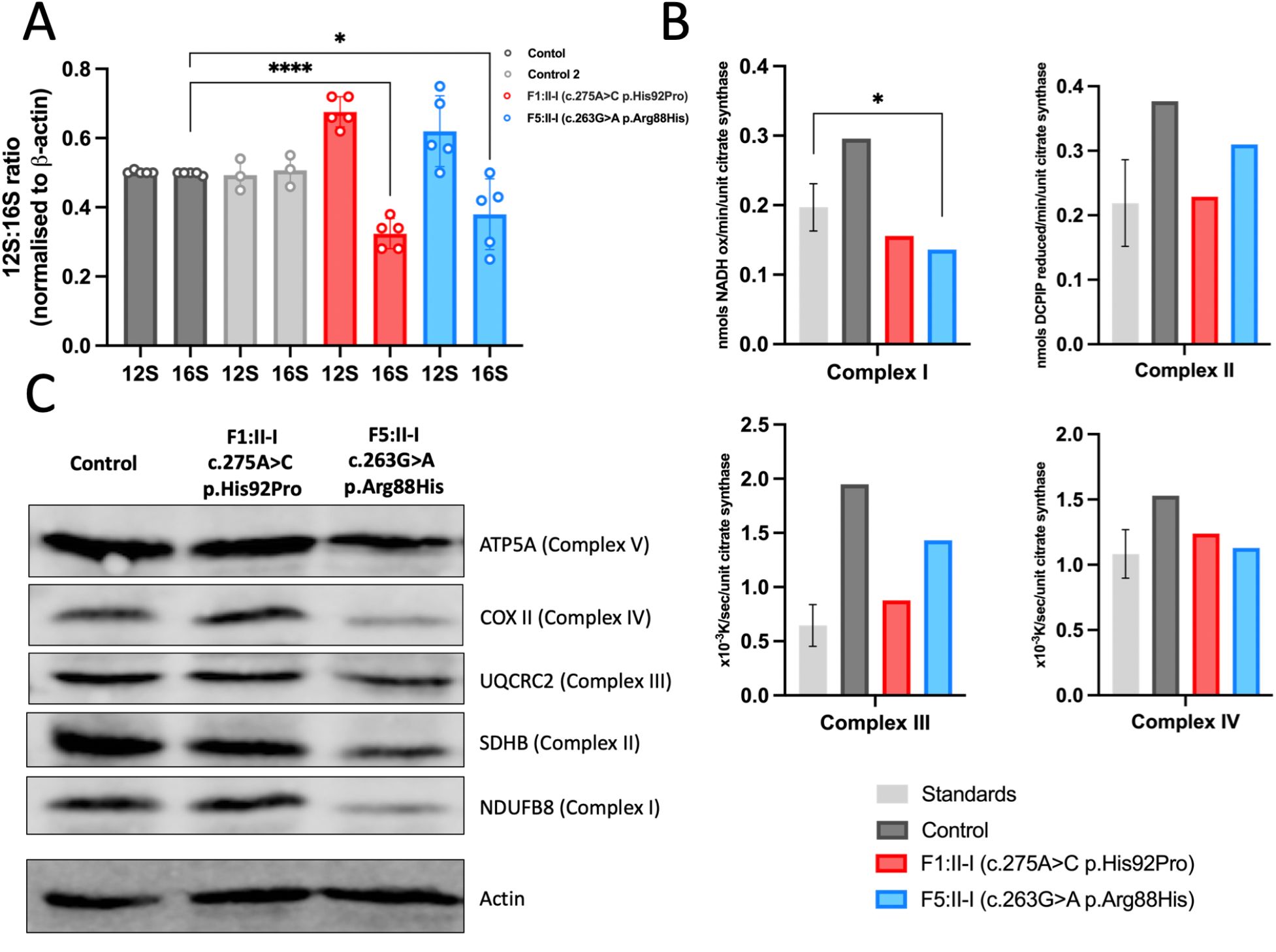
Functional and molecular assays of fibroblasts from affected individuals reveal significant reductions in levels and activities of mitochondrial respiratory chain complexes I and. **IV. (A)** MT-RNR1 (12S) and MT-RNR2 (16S) relative expression levels in fibroblasts from affected individuals and controls. Data expressed as a ratio using relative expression to beta-actin. Error bars represent SD, ****p<0.0001 *p=0.0327, unpaired t-test. **(B)** Mitochondrial respiratory chain enzyme activity assay in control (grey), F1:II-1 (red) and F5:II-1 (blue) fibroblasts. **(C)** Western blot analysis of proteins associated with each of the mitochondrial oxidative phosphorylation complexes in fibroblasts from F1:II-1, F5:II-1 and controls. Image representative of three separate experiments.

To explore more fully the effect of the *MRPL49* missense variants on OXPHOS complexes assembly and the steady-state levels of other mitochondrial proteins, we performed complexome profiling of mitochondria from fibroblasts from affected individuals (Figure 4)^22,23^. Complexome profiling is a quantitative mass-spectrometry (MS) approach previously used to characterize deficiencies of other respiratory complex genes^24–26^. Enriched mitochondrial fractions from both affected individuals and control fibroblasts were subjected to Blue Native electrophoresis (BNE), systematic dissection of the polyacrylamide gel, tryptic digestion, and tandem MS, as reported previously^27,28^. MS data, identification, quantification, and complete interaction profiles of mitochondrial proteins from individuals F1:II-1, F5:II-1 and control fibroblasts were deposited to the ProteomeXchange Consortium^29^ via the PRIDE partner repository^30^ with the dataset identifier (PXD056347). Abundance of the proteins of interest in each discrete section of the BN gel was visualized as heatmaps and line charts (Figure 4). The MS analysis identified all 30 protein components of the mt-SSU and 51 out of 52 protein components of the mt-LSU (Supplemental Figure S4). A few components were expected to be found in their free forms due to their known labile interactions and poor stabilization during BNE^31^ The complexome data reveal a general reduction in the content of mitoribosomes, as evidenced by lower levels of individual mt-LSU and mt-SSU in fibroblasts from both affected individuals compared to controls (Figure 4A). In F1:II-1, we observed a decrease of ∼15% in mt-SSU and 60 % in mt-LSU. In F5:II-1, the decrease was more pronounced, with reductions of ∼40% in mt-SSU and 70% in mt-LSU. Strikingly, variants in *MRPL49* did not change the migration patterns of any of the individual components (Supplemental Figure S4), in particular those from the direct interactors, MRPL4, MRPL15, MRPL57 and MRPL64^32^.

**Figure 4.**
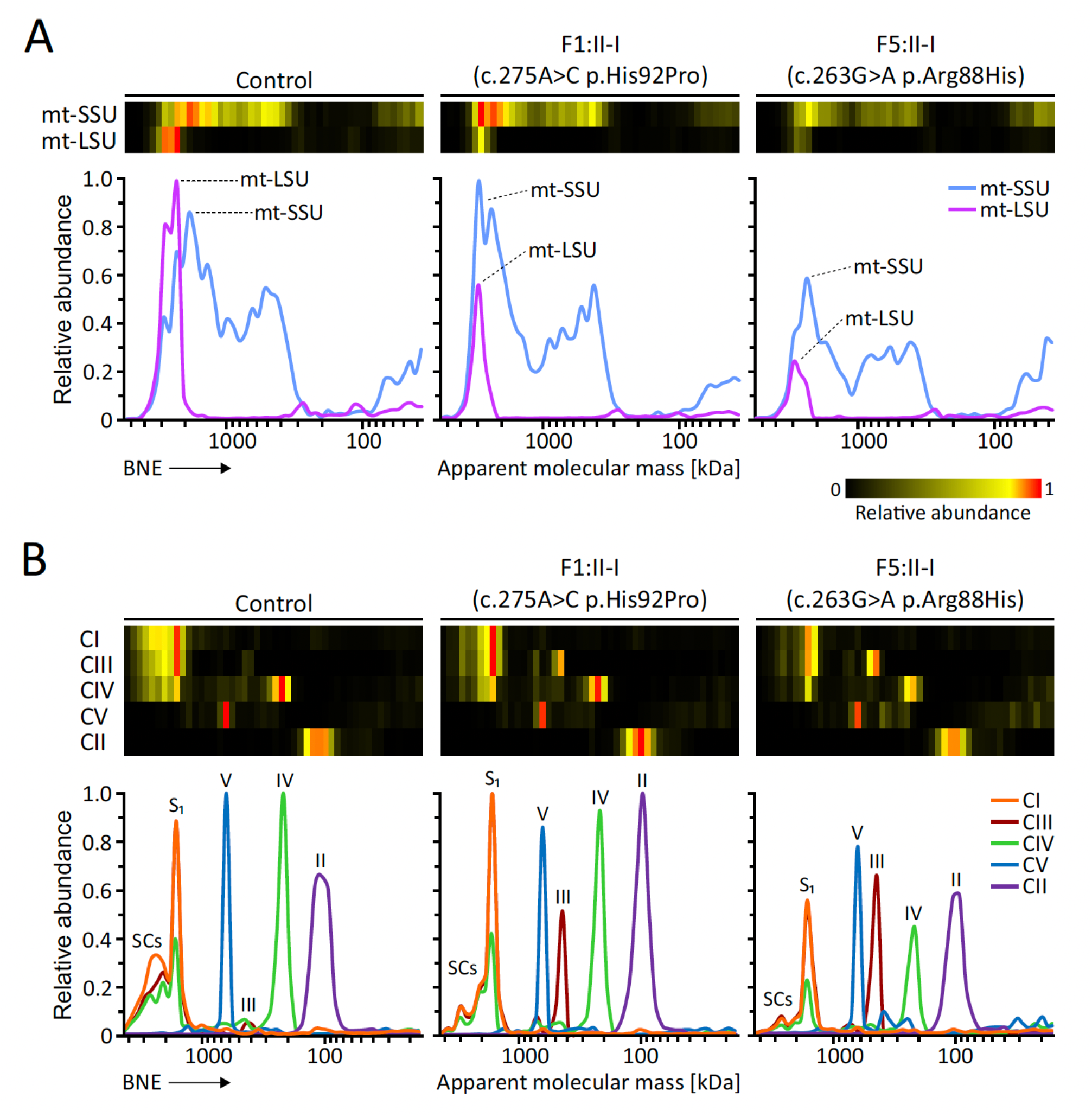
Complexome analysis of mitochondrial ribosomes and OXPHOS complexes from control and F1:II-1 and F5:II-1patient fibroblasts. Enriched mitochondria were solubilized with digitonin, separated by BNE and analyzed by mass spectrometry (MS)-based complexome profiling. Protein abundance profiles of each mitoribosomal subunit and OXPHOS complex were generated by averaging the intensity-based absolute quantification (iBAQ) values of all their individual subunits identified by MS. Resultant profiles are illustrated as heatmaps and 2D profile plots against the apparent molecular mass. **(A)** Abundance profiles of mitoribosomal small (mt-SSU) and large (mt-LSU) subunits showing a visible decrease in the fibroblasts from affected individuals. **(B)** Abundance profiles of the five mitochondrial OXPHOS complexes consistently showing a drop in their content, particularly for complexes I and IV. CI, CII, CIII, CIV and CV stand for complexes I, II, III, IV and V, respectively; S1: respiratory supercomplex formed by complexes I, III and IV; SCs: higher-order respiratory complexes (respirasomes).

The complexome data reflect a reduction in the assembled respiratory supercomplex S1 (I+III+IV) and larger respirasomes (SCs) alongside pronounced reduction in complex I and IV in fibroblasts from the affected individuals (Figure 4B). In F1:II-1 fibroblasts, complex I levels decreased by approximately 30%, while complex IV levels showed a 25% reduction. These alterations are more substantial in F5:II-1 fibroblasts, where complex I and complex IV levels dropped by ∼70% and 65%, respectively. Notably, there were ∼15% and ∼50% reductions in complex III-containing supercomplexes, with corresponding 7- and 11-fold increases in the accumulation of free form complex III in F1:II-1 and F5:II-1, respectively. Complex V levels decreased by ∼20% in both affected individuals. Complex II levels were comparable to the control in F1:II-1, but were ∼15% lower in F5:II-1. Finally, no visible accumulation of assembly intermediates or major defects in OXPHOS biogenesis were observed (Supplemental Figure S5).

Our cumulative data have determined that biallelic variants in *MRPL49* are associated with a complex variable mitochondrial phenotype, characterised by SNHL, POI, leukodystrophy, retinopathy and learning disability. The complexome data detailing marked deficiency of complexes I and IV in fibroblasts from affected individuals are consistent with a reduction in mitochondrial protein translation and define biallelic variants in *MRPL49* as a new cause of COXPD.

MRPL49 is one of a group of proteins within the mt-LSU that has no apparent homolog in bacterial, chloroplast, archaebacterial, or cytosolic ribosomes^2^. However, it does have a homolog in yeast (Img2)^33^, which is required for mitochondrial genome integrity. The specific function of MRPL49 within the mitoribosome has not been determined, but it may compensate for lost rRNA and stabilize bypass segments within the mt-LSU^3^. To date, no *mrpl49* knockout mouse models have been reported^34^. We would predict that such a model would be non-viable. Apart from *Mrpl56* knockout mice^35^, all other knockouts of *Mrpl* genes are non-viable^34^. This prediction of lethality of an *MRPL49* knockout aligns with our data that the disease associated variants are hypomorphic, evidenced by the reduced levels of MRPL49 in complexome profiling of fibroblasts from affected individuals.

To date, variants in over 50 genes have been associated with COXPD^36^ with striking clinical variability of growth retardation, microcephaly, altered tone, leukodystrophy, cardiomyopathy, and liver dysfunction. It is notable that interfamilial phenotypic variability is present even in this small cohort of individuals with *MRPL49* variants. The two families with homozygous His92Pro variants have intrafamilial phenotypic concordance but striking inter-familial differences. The affected females in F1 have classical features of Perrault syndrome, whereas the affected female in F2 has no evidence of ovarian insufficiency and no hearing loss. We undertook immunofluorescence of sections from the ear of wild type adult mice, as previously described^37^ to determine if MRPL49 is present in cells of the inner ear. MRPL49 was observed in the mitochondria in the outer hair cells, inner hair cells and supporting cells, consistent with a role in hearing function, but this pattern does not explain the variable hearing phenotype (Supplemental Figure S2). We infer an undefined genetic modifier underpinning these phenotypic differences.

In contrast to other disorders associated with disruption of the mt-LSU, there was no evidence of cardiomyopathy or liver involvement in individuals with *MRPL49* variants. It will be important to ascertain additional affected individuals to determine the full phenotypic spectrum and the possibility of adult neurocognitive decline as seen in the individual F4 who died at age 36-40 years. However, it is notable that among all the affected individuals reported in this study there is a consistent pattern of progressive leukodystrophy. This consistency contrasts with other genes associated with Perrault syndrome where brain white matter changes are variable in frequency from commonly present in individuals with *CLPP*-associated Perrault syndrome, to absent with *HARS2*-associated disease^13^.

It is notable that the affected individuals had variants altering residues 88 and 92 in MRPL49. These residues lie in a loop of MRPL49 that interacts with the 16S rRNA and variants in this loop result in reduced levels of the 16S rRNA with subsequent effects on mitochondrial protein translation. We would predict that other MRPL49 residues at this interface are likely to be associated with this pleiotropic phenotype. Complexome profiling demonstrated a reduction in levels of all mt-LSU protein components of the mitoribosome in the fibroblasts from affected individuals, which were more pronounced in F5:II-1 (Figure 4, Supplemental Figure S4). A lower but visible drop was also observed in the mt-SSU content (Figure 4, Supplemental Figure S4). Recent cryo-EM studies have indicated that MRPL49 interacts intricately with MRPL4, MRPL15, MRPL57 and MRPL64, alongside the 16S rRNA within the mt-LSU^32^. Given that no changes in the migration patterns of these components were observed, defects at the assembly level seem unlikely. We propose that the two variants in *MRPL49* lead to a loss of the proper three-dimensional architecture of this critical region, resulting in reduced stability of the entire subunit and potential degradation. Consequently, a lower content of fully assembled mitoribosomes likely impacts mitochondrial protein translation rates, diminishing the synthesis of OXPHOS subunits, especially for complex I and complex IV (Figure 4B). Together, these issues compromise not only the correct assembly of OXPHOS complexes but also result in a reduced yield of mitochondrial ATP synthesis, leading to an energy crisis at the cellular level.

In summary, we describe biallelic variants in *MRPL49* associated with a pleiotropic phenotype, including SNHL, POI, leukodystrophy, learning disability and retinopathy in unrelated families. This work expands the number of genes known to cause a phenotype consistent with both Perrault syndrome and COXPD. Further, it increases the known number of genes encoding components of the mt-LSU which are associated with human disease and provides insights into the function of the mitoribosome in the control of mitochondrial homeostasis.

## Data Availability

The MRPL49 variants were submitted to ClinVar (https://www.ncbi.nlm.nih.gov/clinvar/) (GenBank: NM_004927.4; accession numbers SCV004242144 - SCV004242147). The exome datasets supporting this study have not been deposited in a public repository because of ethical restrictions but are available from the corresponding author on request.
The mass spectrometry data have been deposited to the ProteomeXchange Consortium via the PRIDE partner repository with the dataset identifier PXD056347

## Declaration of interest

The authors declare no competing interests.

## Acknowledgements

We thank the families for their participation. This study was supported by the Medical Research Council (MR/W019027/1 RTO, RWT and WGN), Action on Hearing Loss (S35 and S60_Newman); Action Medical Research (GN2494); NIH-National Institute of Child Health and Human Development (NICHD) (R01HD109342 to SML and IS); NIHR Manchester Biomedical Research Centre (IS-BRC-1215-20007 & NIHR203308); the Wellcome Centre for Mitochondrial Research (203105/Z/16/Z to RWT); the UK NHS Highly Specialised “Rare Mitochondrial Disorders of Adults and Children” Service (RWT);The Lily Foundation (RWT), and NIDCD/NIH (R01DC012564 to ZMA) The research team acknowledges the support of the National Institute for Health Research, through the Comprehensive Clinical Research Network.

IW is supported by the Deutsche Forschungsgemeinschaft (DFG): SFB1531-S01 - project number 456687919, TRR267/Z2 - project number 403584255, WI 3728/3-1, project number 515944830. We would like to thank Jana Meisterknecht for her excellent technical support.

## Web Resources

dbSNP, https://www.ncbi.nlm.nih.gov/projects/SNP/ Exome Variant Server, http://evs.gs.washington.edu/EVS/GenBank, https://www.ncbi.nlm.nih.gov/genbank/GeneMatcher, https://genematcher.org/

Ensembl – Variant Effect Predictor, https://www.ensembl.org/Tools/VEP Clustal – Omega, https://www.ebi.ac.uk/jdispatcher/msa/clustalo

GTEx, https://gtexportal.org/home/gnomAD, http://gnomad.broadinstitute.org/FoldX, http://foldxsuite.crg.eu/

LOVD, https://www.lovd.nl/OMIM, https://www.omim.org/

MutationTaster, http://www.mutationtaster.org/PolyPhen-2, http://genetics.bwh.harvard.edu/pph2/

ProteomeXchange Consortium https://www.proteomexchange.org

SIFT, http://sift.bii.a-star.edu.sg

## Data and code availability

The *MRPL49* variants were submitted to ClinVar (https://www.ncbi.nlm.nih.gov/clinvar/) (GenBank: NM_004927.4; accession numbers SCV004242144 - SCV004242147). The exome datasets supporting this study have not been deposited in a public repository because of ethical restrictions but are available from the corresponding author on request.

The mass spectrometry data have been deposited to the ProteomeXchange Consortium via the PRIDE partner repository with the dataset identifier PXD056347 and 10.6019/PXD056347.

### Ethical approval

All individuals provided written informed consent in accordance with local regulations. Ethical approval for this study was granted by the National Health Service Ethics Committee (16/WA/0017) and University of Manchester.

## Notes

### Competing Interest Statement

The authors have declared no competing interest.

### Author Declarations

Ethical approval for this study was granted by the National Health Service Ethics Committee (16/WA/0017) and University of Manchester.

