## Supplemental data for "Biallelic variants in *MRPL49* cause variable clinical presentations, including sensorineural hearing loss, leukodystrophy, and ovarian insufficiency"

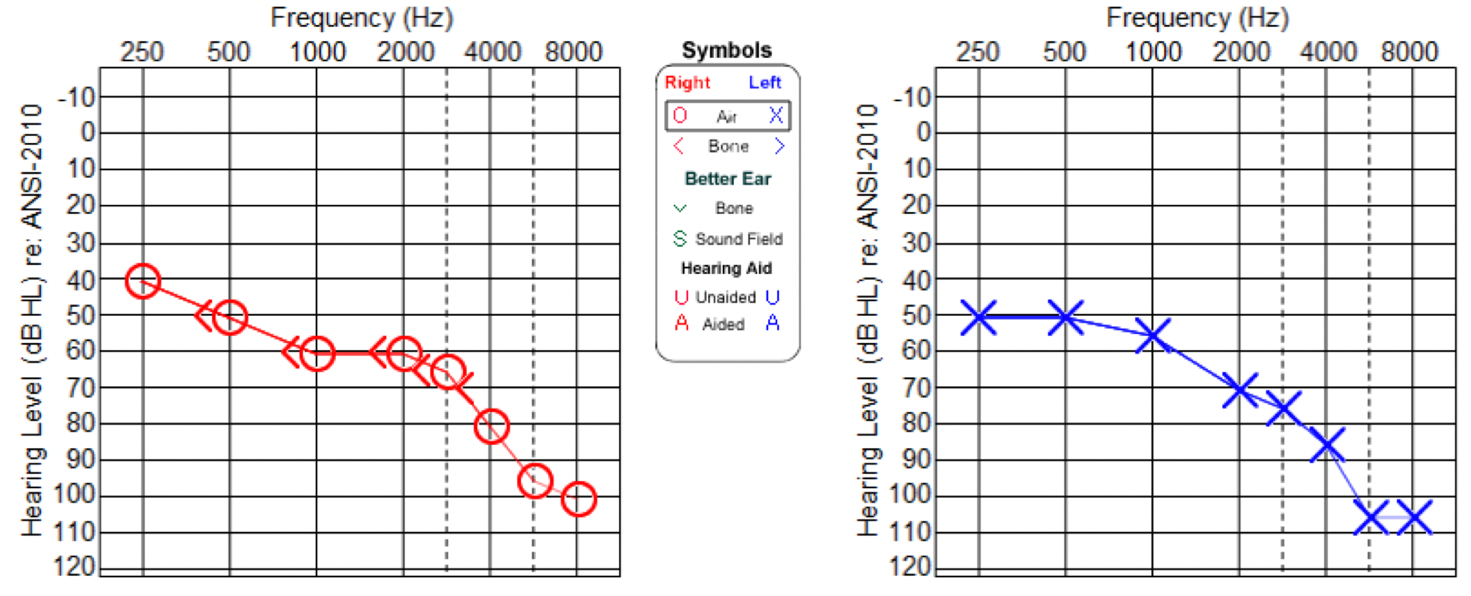

**Figure S1: Audiogram for individual F1:II-1 as an adult.** Measurements reveal a bilateral reduction in hearing levels with increasing frequencies. Measurements for right ear indicated by red circles and left ear indicated by blue crosses.

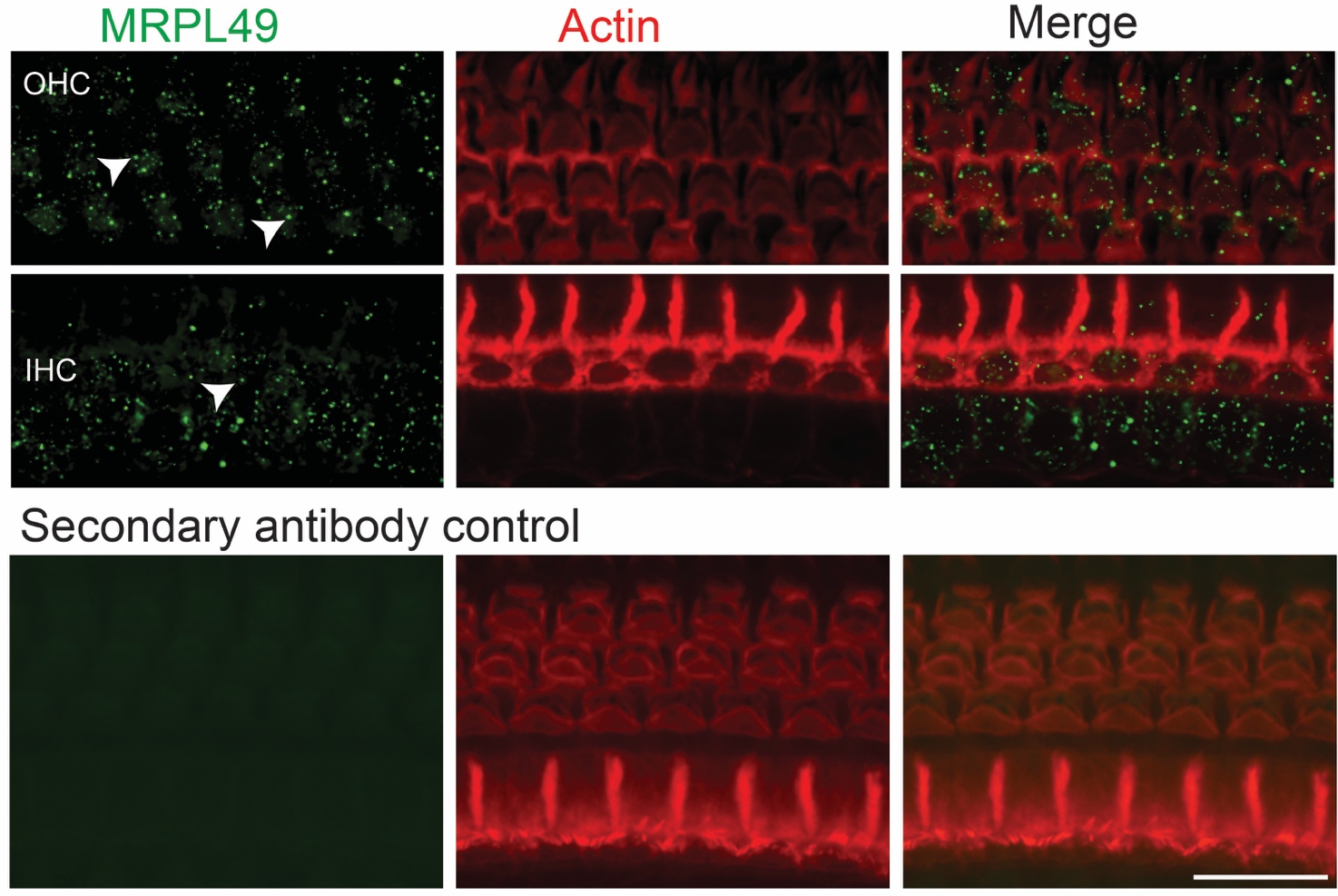
**Figure S2: Whole mount immunostaining of MRPL49 in P12 wildtype mice.** Immunofluorescence of mouse inner ear demonstrated that MRPL49 was localized to mitochondria indicated by arrowheads in outer hair cells, inner hair cells and supporting cells. Scale bar, 20µm.

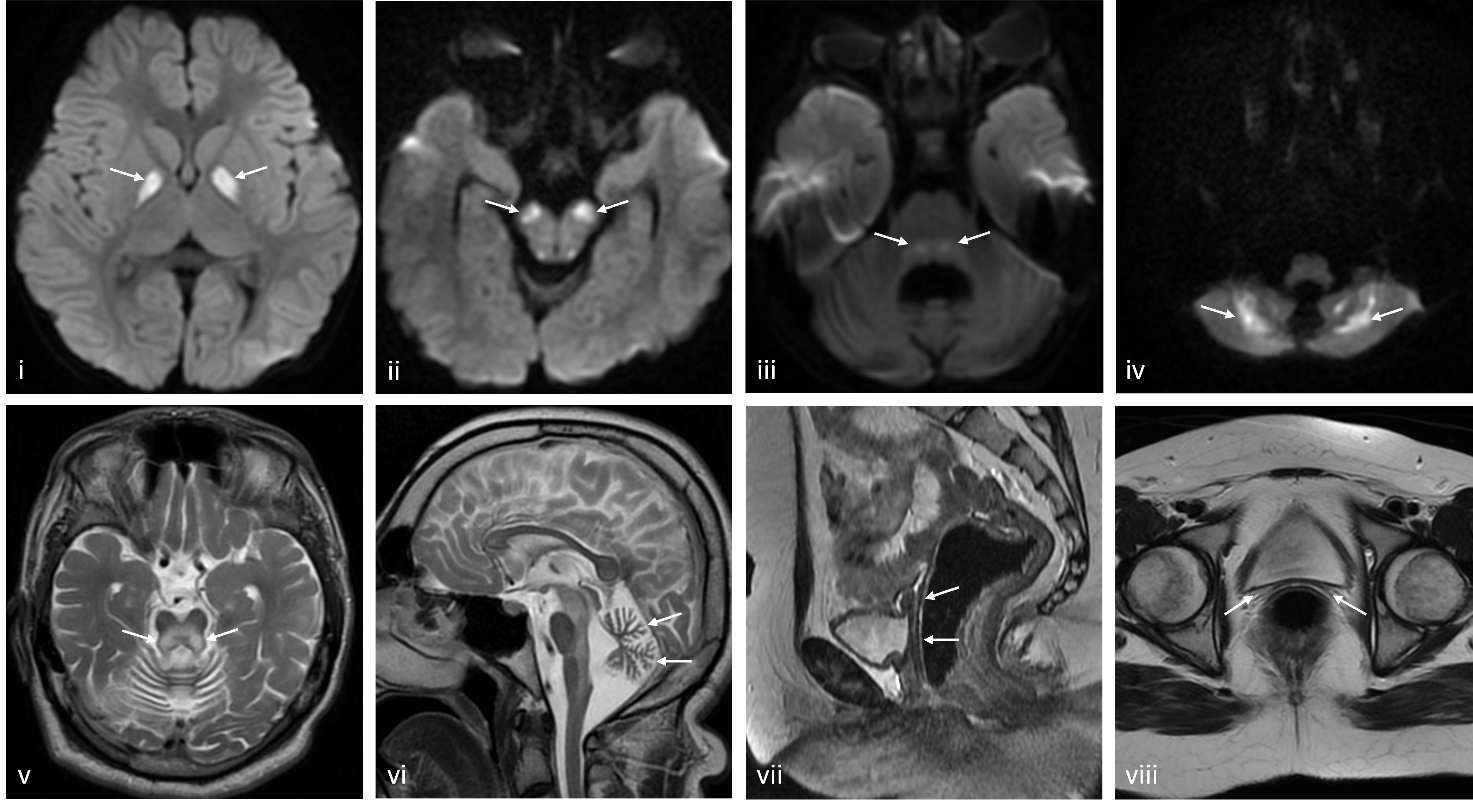

F**igure S3: Further selected MR images.** B1000 diffusion weighted images demonstrate symmetrical diffusion restriction in the globus pallidi (i), substantia nigra (ii), dorsal brainstem (iii) and cerebellum (iv). Selected T2 axial and sagittal images demonstrate dorsal brainstem T2 high signal (v), cerebellar atrophy (vi) and absence of the vagina, uterus and ovaries (vii & viii).

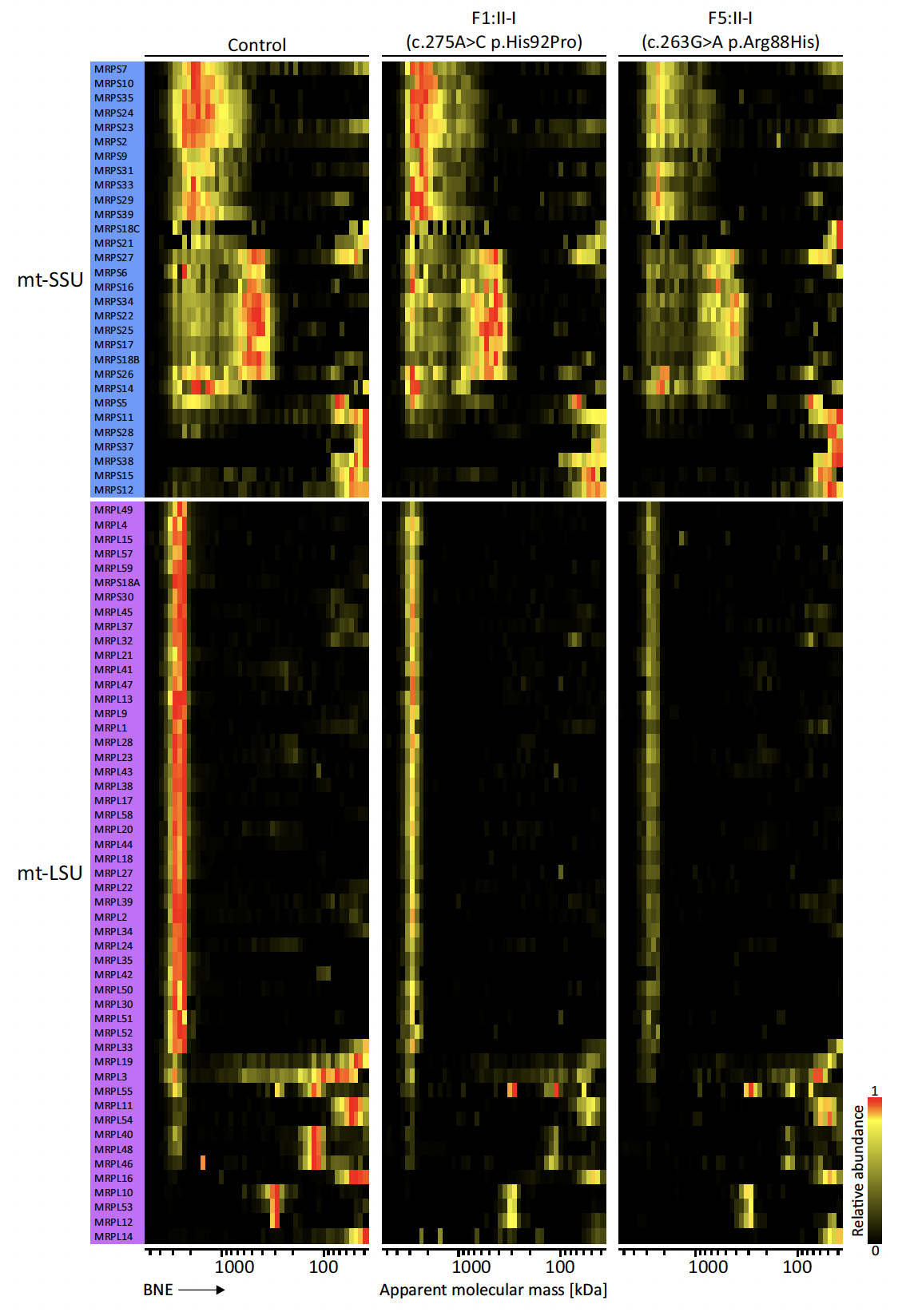

**Figure S4: Expanded complexome profiles of all mitoribosome protein components identified by MS.** Mitochondrial proteins were separated by BNE followed by quantitative mass spectrometry analysis. The iBAQ values of each subunit were normalized using their maximal values across profiles. Resultant relative abundance profiles are shown as heatmaps. Maximum appearance in red, up to 50% in yellow, black indicates that protein abundance is very low, or peptides were not identified in the respective fractions. Mitoribosomal small (mt-SSU) and large (mt-LSU) subunits.

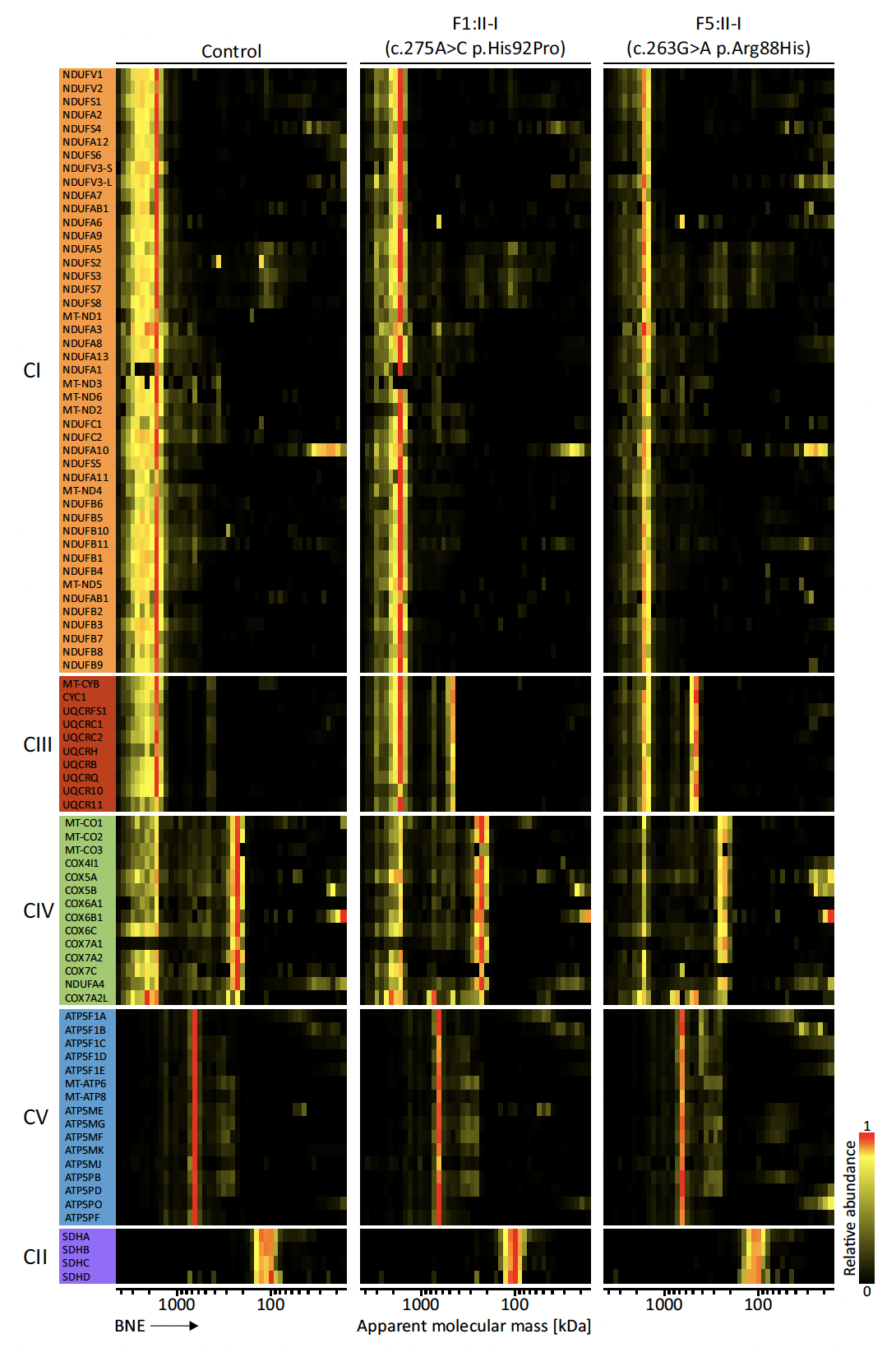

**Figure S5: Expanded complexome profiles of all OXPHOS individual subunits identified by MS.** Mitochondrial proteins were separated by BNE followed by quantitative mass spectrometry analysis. The iBAQ values of each subunit were normalized using their maximal values across profiles. Resultant relative abundance profiles are shown as heatmaps. Maximum appearance in red, up to 50% in yellow, black indicates that protein abundance is very low, or peptides were not identified in the respective fractions. CI, CII, CIII, CIV and CV stand for complexes I, II, III, IV and V, respectively.

| **Gene (alternative name)** | **Length**  **(aa)** | **MIM Gene Reference** | **Phenotype** | **MIM Phenotype Reference** | **No. of families/variants** | **Ref** |
| --- | --- | --- | --- | --- | --- | --- |
| **MRPL1** | **325** | 611821 |  |  |  |  |
| **MRPL2**  **(MRP-L14)**  **(UL2m)** | **305** | 611822 |  |  |  |  |
| **MRPL3** | **348** | 607118 | COXPD9, Cardio myopathy,  SNHL, Leigh Syndrome, | 614582 | Multiple | ^1–3^ |
| **MRPL4** | **311** | 611823 |  |  |  |  |
| **MRPL9** | **267** | 611824 |  |  |  |  |
| **MRPL10** | **261** | 611825 |  |  |  |  |
| **MRPL11** | **192** | 611826 |  |  |  |  |
| **MRPL12** | **198** | 602375 | COXPD45 | 618951 | One | ^4^ |
| **MRPL13** | **178** | 610200 |  |  |  |  |
| **MRPL14** | **145** | 611827 |  |  |  |  |
| **MRPL15** | **296** | 611828 |  |  |  |  |
| **MRPL16** | **251** | 611829 |  |  |  |  |
| **MRPL17** | **175** | 611830 |  |  |  |  |
| **MRPL18** | **180** | 611831 |  |  |  |  |
| **MRPL19** | **292** | 611832 |  |  |  |  |
| **MRPL20** | **149** | 611833 |  |  |  |  |
| **MRPL21** | **205** | 611834 |  |  |  |  |
| **MRPL22** | **206** | 611835 |  |  |  |  |
| **MRPL23** | **153** | 600789 |  |  |  |  |
| **MRPL24** | **216** | 611836 | Cerebellar atrophy, choreoathetosis of limbs and face, intellectual disability, complex I and IV defect |  | One | ^5^ |
| **MRPL27** | **148** | 611837 |  |  |  |  |
| **MRPL28** | **256** | 604853 |  |  |  |  |
| **MRPL30** | **161** | 611838 |  |  |  |  |
| **MRPL32** | **188** | 611839 |  |  |  |  |
| **MRPL33** | **65** | 610059 |  |  |  |  |
| **MRPL34** | **92** | 611840 |  |  |  |  |
| **MRPL35** | **188** | 611841 |  |  |  |  |
| **MRPL36** | **103** | 611842 |  |  |  |  |
| **MRPL37** | **423** | 611843 |  |  |  |  |
| **MRPL38** | **380** | 611844 |  |  |  |  |
| **MRPL39**  **(MRP-L5)** | **338** | 611845 | Early onset mitochondrial disorder |  | Multiple | ^6^ |
| **MRPL40** | **206** | 605089 |  |  |  |  |
| **MRPL41** | **137** | 611846 |  |  |  |  |
| **MRPL42** | **142** | 611847 |  |  |  |  |
| **MRPL43** | **215** | 611848 |  |  |  |  |
| **MRPL44** | **332** | 611849 | Infantile hypertrophic cardiomyopathy  COXPD16 | 615395 | Multiple | ^7–10^ |
| **MRPL45** | **306** | 611850 |  |  |  |  |
| **MRPL46** | **279** | 611851 |  |  |  |  |
| **MRPL47** | **250** | 611852 |  |  |  |  |
| **MRPL48** | **212** | 611853 |  |  |  |  |
| **MRPL49** | **166** | 606866 | POI, SNHL, COXPD |  | Multiple | This report |
| **MRPL50** | **158** | 611854 | POI, SNHL, Heart dysfunction |  | One | ^11^ |
| **MRPL51** | **128** | 611855 |  |  |  |  |
| **MRPL52** | **123** | 611856 |  |  |  |  |
| **MRPL53** | **112** | 611857 |  |  |  |  |
| **MRPL54** | **138** | 611858 |  |  |  |  |
| **MRPL55** | **128** | 611859 |  |  |  |  |
| **MRPL57**  **(MRP63)** | **102** | 611997 |  |  |  |  |
| **MRPL58** | **206** | 603000 |  |  |  |  |
| **MRPL59**  **(GADD45GIP1/ML64)** | **222** | 605162 |  |  |  |  |

**Table S1:** Table listing mt-LSU proteins and their associated disorders

| **Variant** | **c.125_126delTG p.Val42Glyfs*2** | **c.262C>T**  **p.Arg88Cys** | **c.263G>A**  **p.Arg88His** | **c.275A>C**  **p.His92Pro** |
| --- | --- | --- | --- | --- |
| Allele count | 97/1614050 | 49/1613940 | 6/1614036 | 25/1614092 |
| Allele frequency | 6.01e-5 | 3.04e-5 | 3.72e-6 | 1.55e-5 |
| Number of homozygotes | 0 | 0 | 0 | 0 |

**Table S2:** *MRPL49* variant allele frequencies in gnomAD v4.0 (accessed 7/8/24)

|  | **c.262C>T**  **p.Arg88Cys** | **c.263G>A**  **p.Arg88His** | **c.275A>C**  **p.His92Pro** | **c.125_126delTG**  **p.Val42GlyfsTer2** |
| --- | --- | --- | --- | --- |
| Location (hg38) | Chr 11:65125520C>T | Chr 11:65125521G>A | Chr 11:65125533A>C | Chr 11:65124546TG |
| dbSNP | rs758327244 | rs1565337413 | rs770118409 | rs751218133 |
| SIFT | Deleterious  (0.01) | Deleterious  (0) | Deleterious  (0) | - |
| PolyPhen | Benign  (0.338) | Probably damaging  (0.988) | Probably damaging  (0.993) | - |
| CADD | 25.2 | 27.1 | 25.2 | 24.2 |
| Mutation taster | Disease causing  (0.999) | Disease causing  (0.999) | Disease causing  (0.999) | Disease causing  (1) |

**Table S3:**

*In silico* pathogenicity predictions for *MRPL49* variants identified in this study

| **Target** | **Forward sequence 5′-3′** | **Reverse sequence 5′-3′** |
| --- | --- | --- |
| *MT-RNR1* (12s) | TAGAGGAGCCTGTTCTGTAATCGAT | CGACCCTTAAGTTTCATAAGGGCTA |
| *MT-RNR2* (16s) | GCCTGCCCAGTGACACATG | CACGGGCAGGTCAATTTCAC |
| *MRPL49* (Exon 3) | TGGCCCTCTGCAGACTGAGAGC | AGGTCAGGCCAAGAGTCC |
| *MRPL49* (qPCR) | CCGGCTACCAGGATCCCAG | CGGATCACAGTCATCTGCCG |
| *ACTB* | GTGGATCAGCAAGCAGGAGT | GTAACAACGCATCTCATATTTGGAA |

**Table S4:**

Primer sequences used during the study

**Methods:**

**Fibroblast cell culture**

Fibroblast cells were cultured in high glucose Dulbecco′s Modified Eagle′s Medium (Merck) supplemented with 10% (v/v) foetal calf serum (Gibco) and 100U/ml penicillin, 100ug/ml streptomycin, at 37°C with 5% CO_2_ or supplemented with 10% fetal calf serum, 1× non-essential amino acids, 50 U/ml penicillin, 50 μg/ml streptomycin and 50 μg/ml uridine.

**SDS-PAGE and Immunoblotting**

Cells were lysed in RIPA buffer (Sigma) supplemented with protease inhibitor cocktail (Promega), gently agitated at 4°C for 30 minutes and centrifuged at 13,000 rpm for 15 mins. Samples were mixed with SDS-PAGE sample buffer, loaded onto a 12% polyacrylamide gel and run at 180V for up to 90 mins. Proteins were transferred onto a 0.45um PVDF blotting membrane (GE Healthcare) by semi-dry transfer (20V for 30 minutes) before being blocked with TBS-Tween + 5% dried milk power. Primary antibodies specific to each of the mitochondrial respiratory chain complexes as provided by the Total OXPHOS Human WB Antibody Cocktail (ATP5A, UQCRC2, SDHB, COXII, and NDUFB8) (Abcam, ab110411) and Beta-actin (Proteintech; 20536-1-AP, 66009-1-Ig) were incubated overnight at 4°C. Dilutions were 1:500 (OXPHOS cocktail) and 1:5000 (Beta-actin). Membranes were subsequently washed and incubated at room temperature for 90 minutes with either IRDye 680RD Goat anti-Mouse IgG antibody (LI-COR, 926-68070), or IRDye. 800CW Goat anti-Rabbit IgG (LI-COR, 926-32211). Membranes were finally imaged and quantified using the LICOR Odyssey FC imaging system.

**DNA/RNA extraction**

DNA was extracted from patient and control fibroblasts using the DNA minikit following recommended manufacturer’s protocols (Qiagen), DNA was subsequently stored at –80°C.

RNA was extracted from patient and control fibroblasts using Tri-Reagent ® (Merck) following manufacturer’s protocol. RNA was further purified by RNeasy column cleanup (Qiagen) which included an on-column DNase digest step. Purified RNA was immediately used for cDNA synthesis and/or snap frozen at -80°C for storage.

cDNA synthesis: cDNA was generated from purified RNA using GoScript first strand synthesis protocol (NEB) using random hexamer primers (Thermo Fisher Scientific).

**RT-qPCR**

Quantification of patient and control cDNA was performed using PowerUp SYBR Green mastermix (Thermo Fisher) and gene specific primers listed in Table S4. For each separate experiment the mean of triplicate measurements was normalized to ACTB as a housekeeping control.

**Sanger sequencing**

For variant validation in patient fibroblasts, Exon 3 of *MRPL49* was amplified via RT-PCR using “MRPL49 (Exon 3)” primers listed in supplemental table S4. The resulting PCR product was run on a 1% agarose gel, extracted and purified before being prepared for Sanger sequencing. Sanger sequencing was performed by Eurofins Genomics.

**Mitochondrial enzyme activity assay**

Assessment of mitochondrial respiratory chain enzymes in fibroblasts from affected individuals (F1:II-1 and F5:II-1) was performed as previously described^12^

**Complexome analysis**

Cells were homogenized using a motor-driven glass Potter-Elvehjem homogenizer with a Teflon pestle at 2,000 RPM and 40 strokes in a cold room (4 °C). Homogenates were centrifuged for 3 min at 500 x *g* to remove nuclei, cell debris, and unbroken cells. Mitochondrial membranes were sedimented by centrifugation for 10 min at 10,000 x *g*. Protein content was determined using the DC method (Bio-Rad) and mitochondrial enriched pellets (400 µg protein) were resuspended in 40 µl solubilisation buffer (50 mM imidazole pH 7.0, 50 mM NaCl, 1 mM EDTA, 2 mM aminocaproic acid) supplemented with 12 µl 20%(m/v) digitonin (Serva) and processed as described in^13^. Equal protein amounts of samples (100 µg protein) were subjected to Blue Native electrophoresis (BNE) using a 3 to 18% polyacrylamide gradient gel (dimension 14 x 14 cm). After native electrophoresis in a cold chamber, gels were fixed in 50%(v/v) methanol, 10%(v/v) acetic acid, 10 mM ammonium acetate for 30 min and stained with Coomassie Blue dye (0.025% Serva Blue G-250 in 10%(v/v) acetic acid).

Each lane of the native gel was cut into equal fractions and collected in 96-well filter plates. The gel pieces were destained in 60% Methanol, 50 mM ammonium bicarbonate (ABC). Excess solution was removed by centrifugation for 2 min at 600 x *g*. Proteins were reduced in 10 mM DTT, 50 mM ABC for one hour at 56°C and further alkylated for 45 min in 30 mM iodoacetamide. Samples were digested in 5 ng trypsin (sequencing grade, Promega)/μl in 50 mM ABC, 10% acetonitrile (ACN), 0.01% ProteaseMAX surfactant (Promega), 1 mM CaCl_2_ for 16 hours at 37°C. Peptides were eluted with 30% ACN and 3% formic acid (FA), centrifuged into a fresh 96-well PCR plate, dried in a SpeedVac and resolubilized in 1 % ACN, 0.5% FA and stored at -20°C until MS analysis.

The peptides (3 µl of each fraction) were analyzed by LC-MS/MS using a Q Exactive Plus Orbitrap equipped with an UHPLC Dionex Ultimate 3000 instrument (Thermo Fisher Scientific). The peptides were loaded on an Acclaim™ PepMap™ 100 C18 LC Pre-column (0.1 mm x 20 mm, nanoViper, 5 µm, 100 Å) and separated using emitter columns (15 cm length × 100 μm ID × 360 μm OD × 15 μm orifice tip; MS Wil/CoAnn Technologies) filled with ReproSilPur C18-AQ reverse-phase beads of 3 μm, 100 Å (Dr. Maisch GmbH). HPLC and MS specific details are available at the PRIDE entry. MaxQuant 2.0.3.0^14^ was used to perform the proteomic search. The human reference proteome database (UniProt, December 2022, including canonical sequences, isoforms and the two MRPL49 variants) was used for identification with a false discovery rate (FDR) ≤1%. Additional search details are available at the PRIDE entry. To quantify the protein abundances, iBAQ values were calculated. The proteinGroups.txt output file was automatically processed using “process_maxquant” (<https://github.com/joerivstrien/process_maxquant>). The resulting dataset was manually inspected and analyzed using Microsoft Excel. The list of protein groups was hierarchically clustered based on the abundance patterns with an average linkage algorithm (absolute correlation) using Cluster 3.0^15^. Individual profiles were corrected using the sum of iBAQ values of the identified proteins annotated in MitoCarta 3.0^16^ Calibration curves of apparent molecular masses were generated using human membrane and water-soluble protein complexes of known masses. Data were normalized to the maximal iBAQ value across all slices for each protein group to generate the relative abundance profiles and visualized as heatmaps/line charts generated in Microsoft Excel. To analyze multi-protein complexes, additional profiles were generated by averaging the iBAQ values of all individual components identified by MS. Quantification of the abundance differences was done by calculating the area under the curve (AUC) of the regions of interest.

**Mouse inner ear immunostaining:**

Immunofluorescence of mouse inner and outer hair cells was performed as previously described^17^ using an anti-MRPL49 antibody (NBP2-13618 Novus) at 1:200 dilution.
